# Are European Clinical Trial Funders Policies on Clinical Trial Registration and Reporting Improving? – A Cross-Sectional Study

**DOI:** 10.1101/2023.04.05.23288169

**Authors:** Marguerite O’Riordan, Martin Haslberger, Carolina Cruz, Tarik Suljic, Martin Ringsten, Till Bruckner

**Affiliations:** TranspariMED, Bristol, United Kingdom; Aston Medical School, Birmingham, UK; Berlin Institute of Health, Berlin, Germany; University of Guadalajara, Guadalajara, Mexico; Faculty of Medicine, University of Sarajevo, Sarajevo, Bosnia and Hercegovina; Lund University, Lund, Sweden

**Keywords:** Research policy, research funding, clinical trial transparency, publication bias, trial registration, research waste

## Abstract

**Objectives:** Assess the extent to which the clinical trial registration and reporting policies of 25 of the world’s largest public and philanthropic medical research funders meet best practice benchmarks as stipulated by the 2017 WHO Joint Statement,(1) and document changes in the policies and monitoring systems of 19 European funders over the past year.

**Design, Setting, Participants:** Cross sectional study, based on assessments of each funder’s publicly available documentation plus validation of results by funders. Our cohort includes the 25 of the largest public and philanthropic medical research funders in Europe, Oceania, South Asia and Canada. Of these, 19 were previously assessed against the same benchmarks, enabling us to document changes over time.

**Interventions:** Scoring of all 25 funders using an 11-item assessment tool based on WHO best practice benchmarks, grouped into 3 primary categories: trial registries, academic publication and monitoring, plus validation of results by funders.

**Main outcome measures:** The primary outcome measure is how many of the 11 WHO best practice items each of the 25 funders has put into place, and changes in the performance of 19 previously assessed funders over the preceding year.

**Results:** The 25 funders we assessed had put into place an average of 5/11 (49%) WHO best practices. The best practice adopted by most funders 16/25 (64%) was mandating open access publication in journals. In contrast, only 6/25 funders (24%) took PI’s past reporting record into account during grant application reviews. Funders’ performance varied widely from 0/11 to 11/11 WHO best practices adopted. Of the 19 funders for which 2021 baseline data were available,(2) 10/19 (53%) had strengthened their policies over the preceding year.

**Conclusions:** Most medical research funders need to do more to curb research waste and publication bias by strengthening their clinical trial policies.

**Key Points:**

- WHAT IS ALREADY KNOWN ABOUT THIS TOPIC

Strong clinical trial registration and reporting policies coupled with monitoring and sanctions can reduce research waste, curb publication bias and promote transparency. A 2021 assessment found that 19 European medical research funders’ policies fell short of WHO best practices.

- WHAT THIS STUDY ADDS

This is the first study to assess the clinical trial registration and reporting policies of a global cohort of 25 major medical research funders against WHO best practices, identifying gaps in the research waste safeguards of key players across Europe, Oceania, South Asia and Canada. In addition, the study assesses the progress made by 19 funders in the recent past.

- HOW THIS STUDY MIGHT AFFECT RESEARCH, PRACTICE OR POLICY

This study enables funders worldwide to identify and address gaps in their clinical trial transparency policies by pinpointing exactly where they currently fall short of WHO best practices. It also enables policy makers and citizens to assess whether public bodies tasked with furthering medical knowledge have adopted adequate safeguards against research waste and publication bias.

## Introduction

Research waste and publication bias in clinical trials are widespread.(3–5) An estimated 85% of health research is being wasted, with half of all waste due to non-reporting of results alone.(6) Calls to address the problem have a long history.(7) Clinical trials can only inform clinical practice and public health decision-making if and when their results have been made public.(8) However, numerous studies have consistently documented that the results of a significant proportion of clinical trials are never made public.(8) Previous research consistently shows that non-commercial trials have lower publication rates than trials run by industry.(9) Furthermore, trials with ‘positive’ outcomes are more likely to be published, introducing systematic bias into the medical literature.(10) Incomplete reporting of clinical trials wastes taxpayer money and leaves gaps in the scientific record.(11) Current legal and regulatory frameworks provide insufficient safeguards.(12,13)

Since 2013, the World Medical Association’s Declaration of Helsinki has required all clinical trials to be registered and their results to be made public.(8) Public and philanthropic bodies funding clinical trials are uniquely positioned to promote transparency, reduce research waste, and curb publication bias by adopting policies requiring trialists to preregister trials and rapidly make their results public, and monitoring compliance with these rules. The 2017 World Health Organisation (WHO) Joint statement on public disclosure of results from clinical trials (hereafter “WHO Joint Statement”) lists 11 specific policy, monitoring and compliance elements that funders should adopt.(1)

To date, 23 funders and research bodies have formally signed up to the WHO Joint Statement and thereby committed themselves to adopting all 11 elements. Signatories pledged to require grantees to preregister trials on a WHO-linked trial registry, to make trial results public on the same registry within one year of trial completion, to publicly monitor grantees’ compliance with these policies, and to impose sanctions for noncompliance. In May 2022, a World Health Assembly resolution called on funders worldwide to mandate and monitor trial registration and reporting in line with WHO Joint Statement requirements.(8)

Multiple previous studies have assessed funders’ clinical trial policies.(2,11,14) An assessment of 21 European funders conducted in 2021 used 11 items contained in the WHO Joint Statement as its benchmark.(2) It found that funders had only adopted a mean of 4/11 (36%) of WHO best practices in clinical trial transparency. There was a wide variation in performance amongst funders, and some best practice items had been more widely adopted than others. The authors included a template policy document to facilitate the adoption of WHO best practices.(2)

We build on this previous work by assessing a broader cohort of 25 funders worldwide using the same methodology, including 19 of the European funders that had been assessed one year earlier.

## Methods

Study design and reporting were performed in accordance with the Strengthening the Reporting of Observational Studies in Epidemiology (STROBE) reporting guideline for cross-sectional studies.(15)

Our starting point was a cohort of 21 of the largest philanthropic and unilateral public medical research funders in Europe covered by a previous assessment.(2) We removed two funders from that earlier cohort. Bundesministerium für Gesundheit (BMG, the German federal Ministry of Health) was removed because even though it had funded at least one Covid trial in the early stages of the pandemic, it does not routinely directly fund clinical trials. Centre national de la recherche scientifique (CNRS, France) was removed because it does not fund any clinical trials.

We then expanded the cohort of 19 European funders by 6 additional funders to achieve global coverage.(16) We included funders regardless of whether they fund extramural or intramural research, or both. We added two multilateral funders located in Europe that had been excluded from the previous assessment (Horizon Europe and EDCPT), the two major funders in Oceania (NHMRC and HRC), and the largest funder in South Asia (ICMR). We did not include funders in the United States as these were concurrently being assessed by a different study team;(17) to complete coverage of major North American funders, we added Canada’s public funder (CIHR).

We used the same assessment tool and assessment criteria (with minor simplifications) that were used during the 2021 assessment.(2) Scoring of funders was carried out utilising a 11-item assessment tool based on WHO Joint Statement benchmarks.(1) The 11 items fall into 3 broad categories: trial registries (prospective trial registration, registry records kept up to date, results onto registry within 12 months, protocol onto registry within 12 months); academic publication (results published in journal, trial ID included in publications, open access publication); and monitoring and sanctions (investigator’s past reporting record taken into account, trial registration monitored, results reporting monitored, monitoring reports made public). We scored on a YES/NO basis, only awarding points to policy items that fully met the assessment criteria. Policy items that failed to cover all trials, and non-binding ‘supportive’ policy items, were scored as “NO” and earned no points. Possible scores across all items ranged from 0 to 11 points. The protocol, assessment tool, rater guide, adjudication tracker, individual and consolidated score sheets, aggregated data sets, archived funder policies, and correspondence with funders are available on Github.(18)

We searched the websites of all 25 funders during August and September 2022 and filled out a scoring sheet for each funder, capturing relevant policy items. The 6 funders being assessed for the first time were independently assessed by two team members (CC and MR). The remaining 19 funders were assessed by the lead researcher (MOR).

The lead researcher (MOR) then contacted all funders with a copy of the assessment criteria and their scoresheet in November 2022. For funders that had responded to requests to validate the 2021 assessment results, we used the email address of the person who had sent the response in 2021. For all other funders, we used the press department’s email address. One and two weeks after the initial email, a follow-up reminder was sent, copying in the press office where applicable. Funders being assessed for the first time that did not respond to our outreach were re-assessed independently once more by another team member (MH or TS) to ensure that no salient policy elements were overlooked.

The lead researcher (MOR) then compared and merged all assessments into a single consolidated assessment sheet for each funder. As a final quality check, we reviewed the previous year’s assessments of European funders to ensure we had captured all items.

At each stage, a team member not involved in conducting assessments (TB) reviewed all items flagged as uncertain. To ensure comparability of findings across cohorts and time, he determined final scores based on precedents set during two separate studies using the same methodology.(2,17) These decisions were documented in an ‘adjudication tracker’ and have been archived on Github.(18)

## Results

Chart 1 shows the comparative performance of all 25 funders. On average, funders had adopted 5.4/11 WHO best practices in clinical trial transparency (49%).

**Chart 1:**
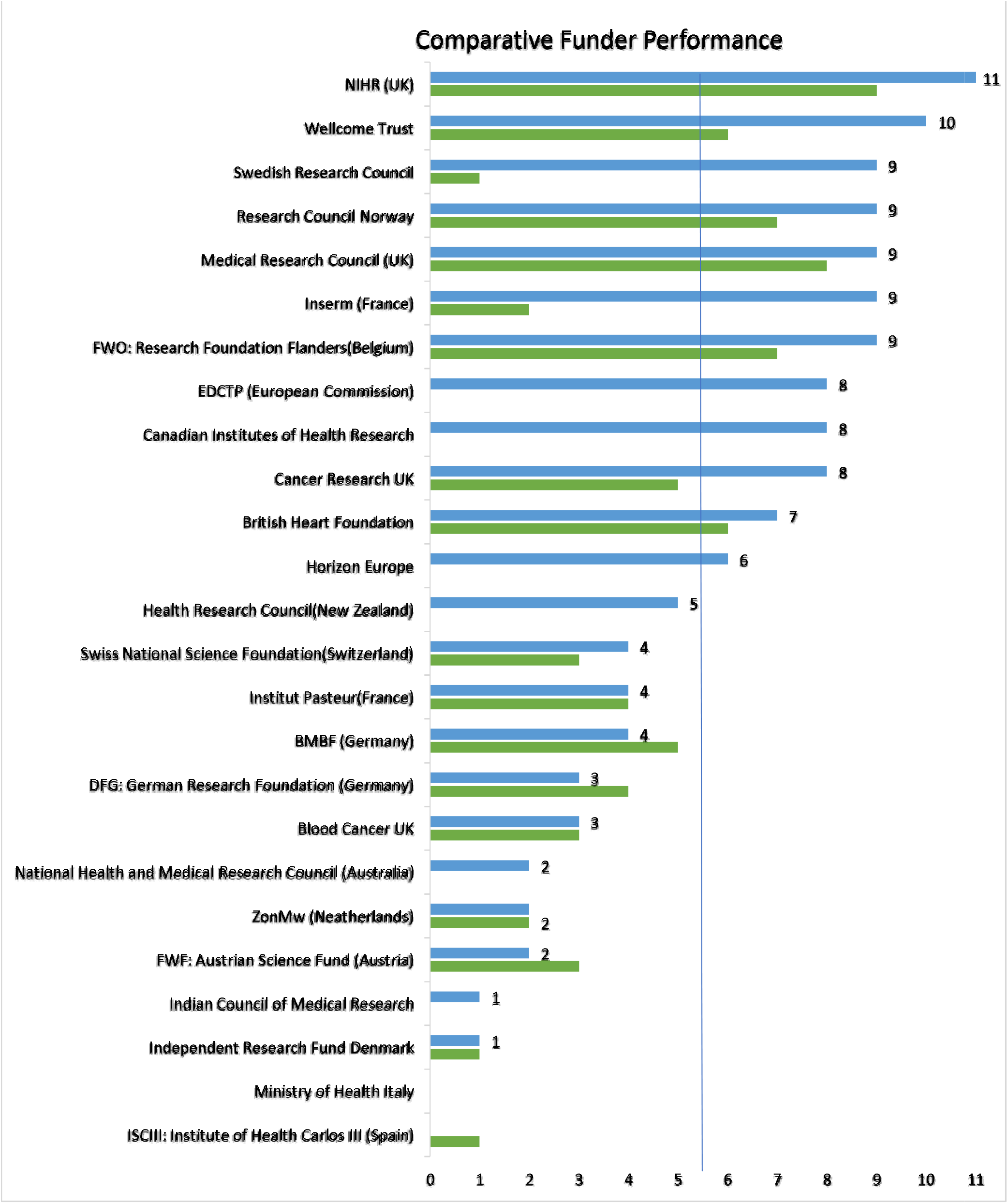
Number of WHO best practices adopted per funder (maximum = 11)

Funders’ performance varied widely. The UK National Institute for Health Research (NIHR) was the only funder that had adopted all 11 policies (100%), followed closely by Wellcome Trust with 10/11 policies (91%). In contrast, Italy’s Ministry of Health and Instituto de Salud Carlos III (ISCIII) both failed to score any points.

Out of the previously assessed European funders, more than half (10/19, 53%) had strengthened their policies over the preceding year, in many cases substantially. The Swedish Research Council and France’s Inserm made the largest gains. The average number of policies adopted by this cohort rose from 4/11 items (36%) to 5.5/11 items (50%) during 2021-2022.

Chart 2 shows which of the 11 policy items had been most widely adopted by the 25 funders in our cohort.

**Chart 2:**
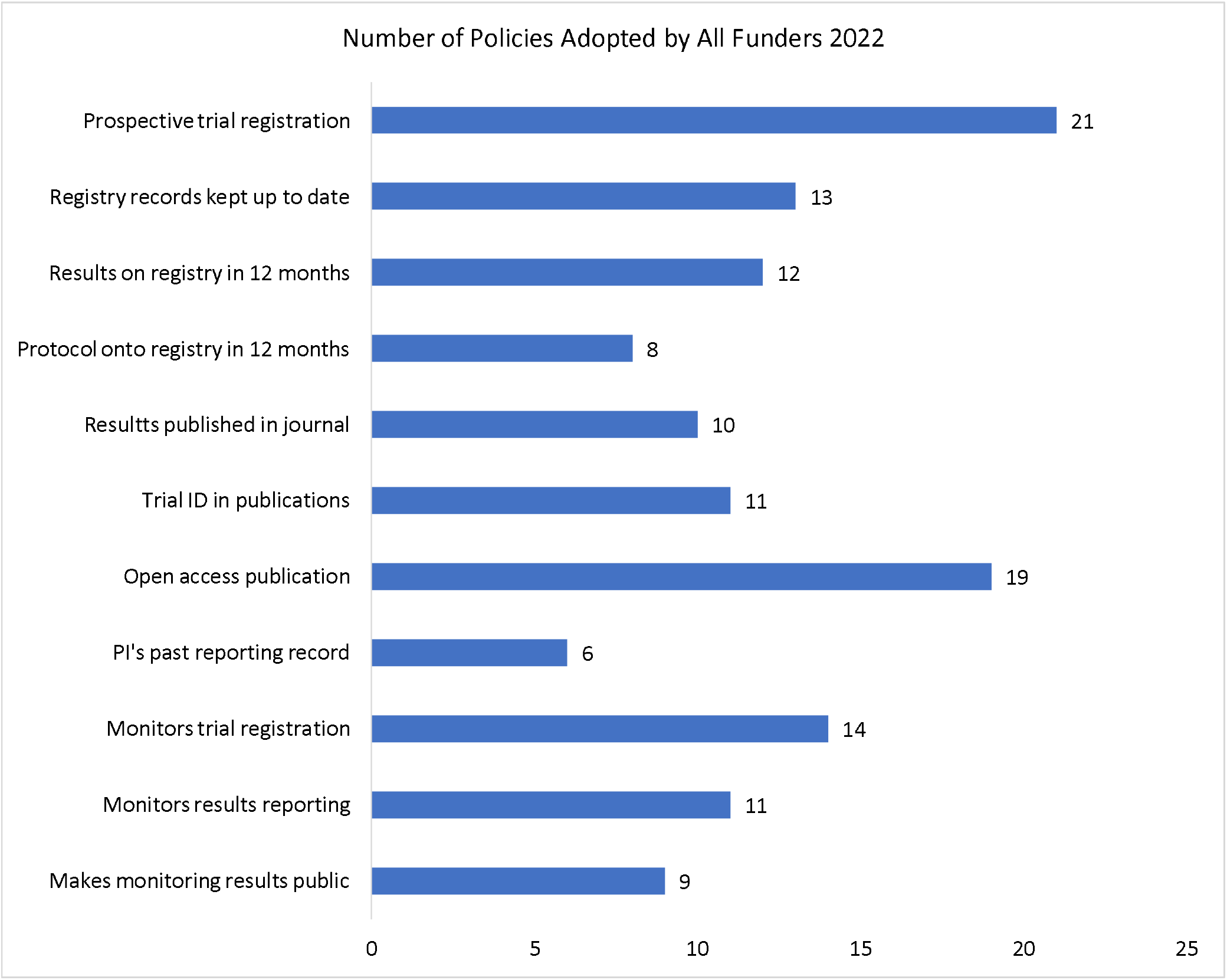
Number of funders adopting specific policy items (maximum = 25)

Prospective trial registration, which has been a global ethics requirement since 2008, was the most widely adopted policy (21/25 funders, 84%), followed by open access publication (19/25 funders, 76%).(8) In contrast, less than a third of funders (8/25 funders, 32%) require their grantees to make their trial protocols publicly available on registries.

Nearly half of funders (12/25 funders, 48%) require trial results to be made public on registries within a year of trial completion, which is a key mechanism for speeding up the disclosure of research outcomes.

A majority of funders (14/25 funders, 56%) monitor whether grantees register trials and make results public. However, when deciding whether to award new grants, fewer funders (9/25 funders, 36%) take into account whether applicants have made trial results public in the past.

Chart 3 shows which policy items the 19 previously assessed European funders added during 2021-2022.

**Chart 3:**
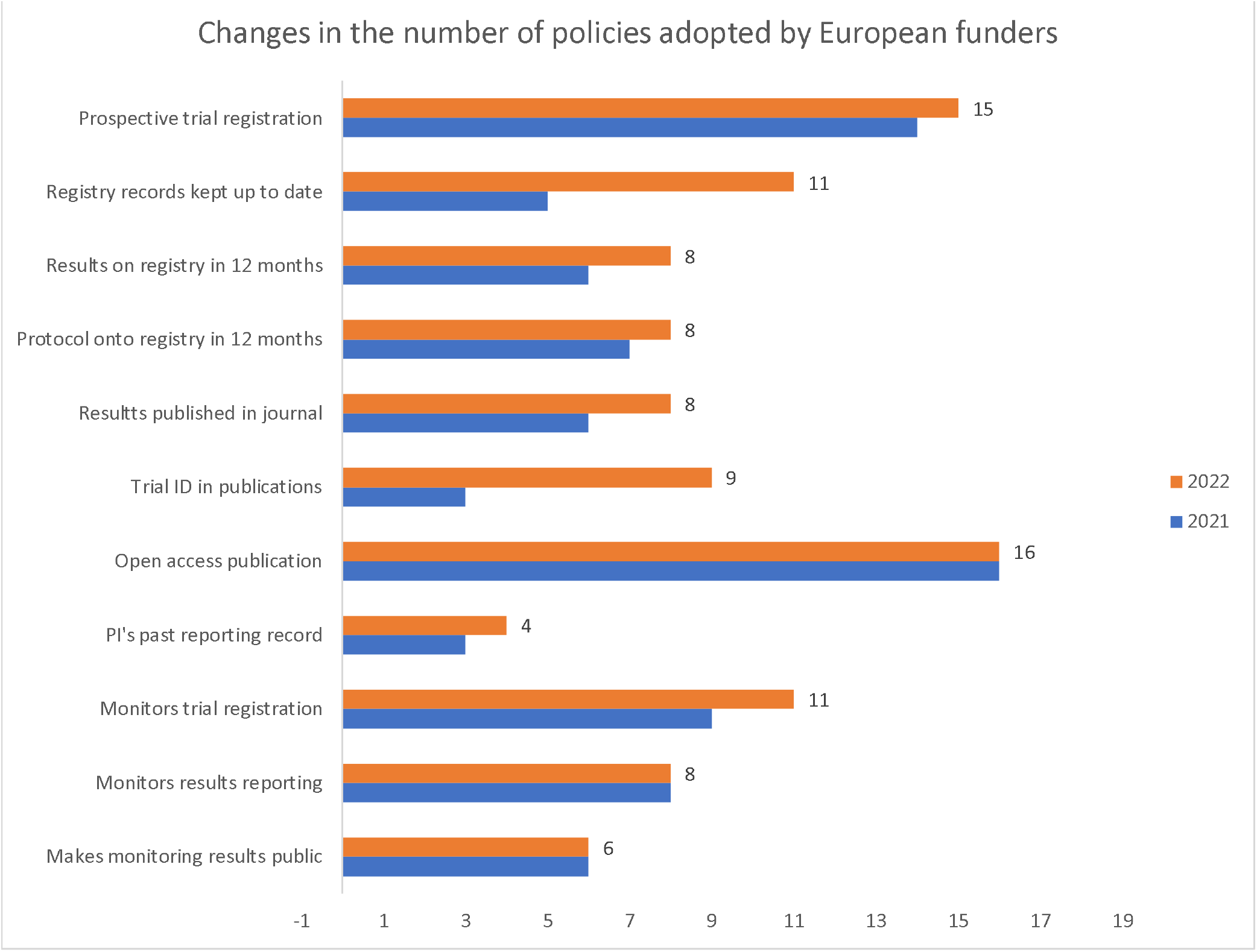
Policy items added by 19 European funders during 2021-2022.

Funders adopted new policy items across the whole range of 11 WHO best practices. The only exception was open access publication, for which the baseline was already very high; note that open access policies tend to be set at the wider institutional level rather than specifically for clinical research.

For other policy items, growth in uptake among funders was uneven. The most frequently added new policy items were inclusion of clinical trial registry ID numbers in publications and requirements to keep registry records up to date; each of these items was adopted by 6 additional funders. Of note, 3 funders initiated compliance monitoring activities during 2021-2022.

Funders’ efforts to strengthen their policies appear to sometimes have been ad hoc rather than systematic. For example, 11 funders now require grantees to keep registry records up to date, a task that requires diligence from trialists throughout the life cycle of a trial. In contrast, only 11 funders require grantees to include trial ID numbers in publications, which is a very simple action that activity typically needs to be performed only once after the end of a trial.

Table 1 provides a granular overview of the individual policy items adopted by each of the 25 funders. We include this table to enable funders to identify and address remaining gaps in their policies.

**Table 1:** Policy items adopted by each funder and remaining gaps.

|  | Prospective trail registration | Registry records kept up to date | Results onto registry in 12 months | Protocol onto registry in 12 months | Results published in journal | Trial ID in publications | Open access publications | PI'S past reporting record | Monitors trial registration | Monitors results reporting | Makes monitoring reports public |
| --- | --- | --- | --- | --- | --- | --- | --- | --- | --- | --- | --- |
| BCUK | YES | NO | NO | NO | YES | NO | YES | NO | NO | NO* | NO |
| BHF | YES | YES | YES | YES | NO* | YES | YES | NO | YES | NO | NO |
| BMBF | YES | YES | YES | NO | YES | NO | NO* | NO | NO | NO | NO |
| CRUK | YES | YES | YES | NO | NO* | YES | YES | NO | YES | YES | YES |
| DFG | YES | NO | NO | NO* | NO* | NO | YES | NO | YES | NO | NO |
| FWF | YES | NO | NO | NO | NO | NO | YES | NO | NO | NO | NO |
| FWO | YES | YES | YES | NO | YES | YES | YES | YES | YES | YES | NO |
| IRFD | NO | NO | NO | NO | NO | NO | YES | NO | NO | NO | NO |
| INSERM | YES | YES | YES | YES | YES | YES | YES | NO | YES | YES | NO |
| IP | YES | YES | NO | NO | YES | NO | YES | NO | NO | NO | NO |
| ISCIII | NO | NO | NO | NO | NO | NO | NO | NO | NO | NO | NO |
| ITALY MOH | NO | NO | NO | NO | NO | NO | NO | NO | NO | NO | NO |
| MRC | YES | YES | NO | YES | NO* | YES | YES | YES | YES | YES | YES |
| NIHR | YES | YES | YES | YES | YES | YES | YES | YES | YES | YES | YES |
| RCN | YES | YES | NO | YES | YES | YES | YES | NO | YES | YES | YES |
| SNSF | YES | NO | NO | YES | NO* | NO | YES | NO | YES | NO | NO |
| SRC | YES | YES | YES | YES | NO* | YES | YES | NO | YES | YES | YES |
| WELLCOME TRUST | YES | YES | YES | YES | NO | YES | YES | YES | YES | YES | YES |
| ZonMw | NO* | NO | NO | NO | YES | NO | YES | NO | NO | NO | NO |
| CIHR | YES | NO | YES | NO | YES | YES | YES | NO | YES | YES | YES |
| EDCTP | YES | YES | YES | NO | YES | NO | YES | YES | YES | YES | NO |
| HORIZON EU | YES | YES | YES | NO | NO* | YES | NO# | YES | NO | NO | YES |
| HRC | YES | NO | YES | NO | NO* | NO | NO* | NO | YES | YES | YES |
| ICMR | YES | NO | NO | NO | NO* | NO | NO | NO | NO | NO | NO |
| NHMRC | YES | NO | NO | NO | NO | NO | YES | NO | NO | NO | NO |

The “YES” fields denote mandatory policy items that apply to all clinical trials. The “NO” fields denote the complete absence of a policy item.

In 15 instances, funders encouraged a practice but did not make it compulsory. Of note, 9 funders encouraged results to be published in journals, but did not make this compulsory. In 1 instance, the scope of a policy item was limited to drug trials only. Non-binding policies, where a funder encourages a practice but does not mandate it, are marked with “NO*” below. Non-comprehensive policies that apply to only some types of trials are marked with “NO#”

An overview of nonbinding and non-comprehensive policies is provided in the Supplement.

### Strengths and Limitations

This is the first global assessment of medical research funders’ clinical trial policies that is fully based on WHO best practice benchmarks. Multiple independent ratings, review and consolidation by a third researcher, transparent adjudication, and respondent validation strengthened data quality and reliability. Archiving of all project tools and documentation on GitHub enables independent replication, including with other cohorts. A visual aid (Table 1) enables funders to easily identify gaps in their policies. Funders can refer to a previously published template policy document for additional guidance.(2)

Our study has two limitations. Out of 25 funders, 11 funders did not respond to our outreach despite repeated efforts and a deadline extension. As a result, relevant policy items for those funders may have been missed, especially if these were not publicly accessible online. For example, the Swiss National Science Foundation noted that although our assessment accurately reflected publicly available information, they had additional requirements that were not openly accessible.

The second limitation is that funder policies do not necessarily translate into improvements in actual practice, especially if funders do not actively monitor grantees’ compliance with their requirements.(19,20)Our study provides a useful starting point for other researchers to assess to what degree and under what conditions funder policies influence clinical trial registration and reporting in practice.

## Discussion

Our study shows that medical research funders vary widely in their adoption of WHO best practices. Even though 15/25 (60%) funders in our cohort have formally committed to adopting all 11 policy items by signing up to the WHO Joint Statement, only a single funder in the cohort has fully delivered on its promise so far. On the positive side, several funders have significantly strengthened their policies over the past year, and a third of funders in our cohort have by now put into place 9 or more of the 11 WHO policy items. While many funders still need to do more to curb research waste and publication bias, the trend is clearly positive. We plan to re-assess all funders in future to document further improvements.

We also found large variations in the frequency with which individual policy items had been adopted by funders. The adoption of open access policies is now extremely widespread. We found that many funders set out formal requirements without monitoring grantee compliance. A negative surprise was that 4 funders still do not require all clinical trials to be registered, even though trial registration is both a long-standing global ethics requirement and a precondition for publication in a respectable journal as well as WHO best practice.(8,21)

Our data indicate that existing research waste safeguards could often be strengthened at no cost to the funder itself, and at negligible cost to grantees. In some cases, funders require grantees to perform time-intensive tasks without requiring related simple tasks to be concurrently performed. For example, several funders require summary results to be uploaded onto trial registries, but do not require grantees to upload study protocols at the same time, which could be done within a few minutes. Other funders mandate journal publication, but do not require grantees to copy and paste trial ID numbers into their scientific papers. Table 1 can help funders to identify such potential easy wins.

## Conclusion

The UK’s National Institute for Health Research has fully adopted all WHO best practices in clinical trial transparency. Several other funders have also put strong research waste safeguards in place. While many funders’ policies still fall significantly short of WHO best practices, average funder performance appears to be improving.

Each of the 11 WHO best practices has been adopted by at least 8 funders in our cohort, demonstrating feasibility. As the WHO has noted, the resource allocation, public health and scientific benefits of rapid and comprehensive outcome reporting far outweigh the modest implementation costs.(1) Funders’ experiences to date show that effective research waste safeguards can be put into place without antagonizing grantees or burdening them with excessive red tape.(14,22–24)

We urge funders to further strengthen their policies, and to concurrently support and adequately compensate their grantees’ trial registry management and outcome reporting efforts. Recent experience shows that strong funder policies alone are insufficient to prevent research waste, so it is essential for funders to monitor grantees’ compliance.(20) We encourage funders to use such monitoring data to identify and highlight strong as well as weak performers.

## Data Availability

All data produced in the present study are available upon reasonable request to the authors

https://github.com/mriordan7/Consilium-Scientific-Project/blob/main/README.md

## Footnotes

### CONTRIBUTORS

MOR coordinated the study team, managed communications with funders, analysed the data, and wrote the manuscript. MOR, MR, CC, MH and TS contributed to acquisition of data. TB developed the study protocol, acted as adjudicator, and edited the manuscript. All authors reviewed the manuscript.

### PATIENT CONSENT FOR PUBLICATION

Not required.

### ETHICS APPROVAL

NHS REC ethics waiver obtained on 18 October 2022.

### DATA AVAILABILITY STATEMENT

All data relevant to the study are included in the article or uploaded onto Github as supplementary information.

## Abbreviations

BCUK: Blood Cancer UK
BHF: British Heart Foundation
BMBF: Federal Ministry of Education and Research
CRUK: Cancer Research UK
DFG: Deutsche Forschungsgemeinschaft
FWF: Der Wissenschaftsfonds
FWO: Research Foundation Flanders
IRFD: Independent Research Fund Denmark
INSERM: Institut Nationale de la sante et de la recherche medicate
IP: Institut Pasteur
ISCIII: Instituto de Salud Carlos III ITALY
MOH: Ministry of Health of Italy
MRC: Medical Research Council
NIHR: National Institute for Health Research
RCN: Research Council Norway
SNSF: Swiss National Science Foundation
SRC: Swedish Research Council
ZonMw: THE NETHERLANDS ORGANISATION FOR HEALTH RESEARCH AND DEVELOPMENT
CIHR: The Canadian Institutes of Health Research
EDCTP: European & Developing Countries Clinical Trials Partnership (EDCTP) [via the European Commission]
HRC: Health Research Council of New Zealand
ICMR: Indian Council of Medical Research
NHMRC: National Health and Medical Research Council (Australia)

